# Phenome-wide comorbidity network analysis reveals clinical risk patterns in enthesopathy and enthesitis

**DOI:** 10.1101/2025.04.21.25326169

**Authors:** Yonghyun Nam, Dong-gi Lee, Jakob Woerner, Se-Hwan Lee, Min Ji Lee, Sung-Han Jo, Jaeun Jung, Su Chin Heo, Chris Hyunchul Jo, Dokyoon Kim

## Abstract

**Background:** Enthesopathy and enthesitis, including rotator cuff disease and other tendon disorders, represent a heterogeneous group of musculoskeletal conditions with complex etiologies. Understanding how systemic health profiles influence their onset remains a critical challenge in musculoskeletal medicine.

**Methods:** We conducted a large-scale, phenome-wide comorbidity analysis using longitudinal electronic health records (EHR) from 432,757 UK Biobank participants. Incident cases of peripheral enthesopathies were compared to controls across 434 baseline disease phenotypes. A directed ego network was constructed to link significantly associated comorbidities to the target condition using odds ratio-based associations. Unsupervised clustering via UMAP and DBSCAN identified data-driven comorbidity clusters, which were consolidated into unified endotypes-interpreted as distinct systemic profiles contributing to disease risk. Additionally, metapath-based trajectory analysis was applied to uncover temporally structured multimorbidity chains leading to disease onset.

**Results:** We identified 183 baseline conditions significantly associated with the future development of enthesopathy (FDR < 0.05). Network clustering revealed eight comorbidity clusters, which were consolidated into four unified endotypes: Metabolic-Psychosomatic, Inflammatory-Multisystem, Mechanical-Injury-driven, and Aging-Intervention-related. Metapath analysis uncovered common three-step disease trajectories, such as metabolic-infectious-musculoskeletal and inflammatory skin-to-joint progressions, highlighting potential mechanistic pathways. These endotypes showed diverse clinical features but shared biological coherence, suggesting that different systemic health profiles can converge to drive tendon-related disease.

**Conclusions:** This study introduces a scalable framework for identifying systemic multimorbidity patterns underlying enthesopathy and enthesitis using phenome-wide comorbidity networks. By integrating network clustering and metapath analysis, we uncover interpretable, data-driven endotypes that may inform individualized risk assessment and targeted care strategies. These findings contribute to the growing field of biobank-scale disease modeling and offer a foundation for precision approaches in musculoskeletal medicine.

## 1. Introduction

Enthesopathy and enthesitis, including tendinopathies and partial-to full-thickness rotator cuff tears, are among the most common musculoskeletal disorders contributing to chronic pain, disability, and healthcare burden worldwide^1,2^. These disorders affect the enthesis, the connective tissue interface between tendons or ligaments and bone, encompassing a heterogeneous group of conditions with different etiologies^3,4^. Underlying mechanisms include mechanical overload, systemic inflammation, metabolic dysregulation, and age-related degeneration^5-7^. This clinical and biological heterogeneity presents major challenges for early diagnosis, prognostic assessment, and the design of personalized therapeutic strategies ^8^. Recent advances in tendon biology have revealed microenvironmental factors that drive disease progression, including stem cell exhaustion, chronic inflammation, and impaired healing. Despite these insights, most current treatments remain symptomatic rather than mechanistic. Common interventions such as corticosteroid injections, physical therapy, or surgery often fail to address the underlying pathobiology, leading to persistent pain and high rates of re-tear in rotator cuff disease.

Recent evidence suggests that coexisting conditions, particularly those identifiable through longitudinal health records, may offer valuable insight into the pathways leading to enthesopathy and enthesitis^9,10^. While prior studies have examined isolated risk factors such as obesity, diabetes, or smoking^11-13^, few have systematically explored how broad comorbidity profiles may reveal underlying systemic contributors or stratify patient subgroups. In this context, phenome-wide comorbidity analysis provides a powerful approach for capturing the full spectrum of diseases that precede or co-develop with the target condition^14-16^. By evaluating all diagnostic phenotypes simultaneously, this method enables discovery of both expected and previously unrecognized associations, revealing how enthesopathy and enthesitis may arise within distinct clinical trajectories shaped by metabolic, inflammatory, neurological, or psychosocial factors. These distinct trajectories suggest that the disorder does not arise from a single pathogenic pathway, but rather from multiple systemic contexts, each potentially requiring different treatment approaches. Given this heterogeneity, there is increasing recognition that shoulder tendon disease consists of biologically distinct subgroups, or endotypes, each with different risk trajectories. These limitations underscore the need for personalized approaches beyond generalized use of corticosteroids or physical therapy. Identifying endotypes through data-driven analysis of systemic comorbidity patterns could support more precise and mechanism-informed models of care.

To address this gap, we performed a large-scale phenome-wide comorbidity analysis of peripheral enthesopathies and allied syndromes using longitudinal electronic health record (EHR) data from 432,757 participants in the UK Biobank. We focused on incident cases, defined as individuals who developed the condition after enrollment, and evaluated 434 baseline phenotypes for associations with future onset of enthesopathy and enthesitis. Comorbidities significantly enriched among these individuals were then used to construct a directed ego–alter disease network centered on the target phenotype. Using unsupervised network clustering and metapath-based trajectory analysis, we identified distinct multimorbidity patterns that may represent unique systemic contexts contributing to the pathogenesis of enthesopathy and enthesitis. This network-guided framework offers a scalable and interpretable strategy to characterize heterogeneity in musculoskeletal disorders and sets the stage for endotype-informed risk stratification and precision intervention.

## 2. Results

To investigate systemic risk patterns associated with incident enthesopathy and enthesitis, we conducted a multistep analysis integrating phenome-wide association testing, network modeling, unsupervised clustering, and trajectory analysis. Starting with 434 baseline phenotypes derived from hospital diagnosis data, we identified 183 comorbid conditions significantly associated with future onset of the target disease. These associations were used to construct a directed ego–alter disease network, which revealed eight distinct comorbidity clusters through graph-based clustering. To enhance interpretability, we grouped the eight comorbidity clusters into four unified endotypes based on shared clinical patterns and biological plausibility. We then applied a metapath framework to trace frequently observed diagnostic sequences leading to enthesopathy and enthesitis, uncovering interpretable multimorbidity trajectories.

### 2.1 Study Population

We analyzed longitudinal electronic health record (EHR) data from 432,757 participants in the UK Biobank who had complete baseline data and follow-up information. To focus on disease onset, we restricted the analysis to incident cases of peripheral enthesopathies and allied syndromes (phecode 726), defined as first diagnosis occurring after the baseline enrollment date. Individuals with diagnoses prior to baseline were excluded to ensure clear temporal sequencing. Case-control status was defined as follows: cases were individuals newly diagnosed with peripheral enthesopathies and allied syndromes after baseline (N=14,515; 3.4%), while controls were those without any diagnosis of this condition throughout the observation period (N=418,242; 96.6%). This phenotyping approach enabled assessment of pre-existing health conditions prior to disease onset in a temporally consistent manner. The median age at enrollment was 58 years (interquartile range: 51–64), and 55% of participants were female. To evaluate pre-existing health status, we examined 434 diagnostic codes representing baseline disease history across 16 major clinical categories, including injuries and poisonings, endocrine and metabolic disorders, circulatory system diseases, neurological conditions, mental health disorders, and others. These diagnoses represent underlying conditions for subsequent comorbidity and network-based analyses.

### 2.2. Phenome-wide Comorbidity Analysis Identifies Diverse Risk Profiles

To identify underlying conditions associated with the future development of enthesopathy and enthesitis, we conducted a phenome-wide comorbidity analysis using diagnostic histories recorded prior to baseline. For each of the 434 candidate conditions, we constructed a 2×2 contingency table comparing its presence or absence before baseline enrollment with the incidence of enthesopathy and enthesitis after enrollment. Odds ratios were calculated to quantify the strength of association, and statistical significance was assessed using Fisher’s exact test. Multiple testing correction was applied using the false discovery rate (FDR).

A total of 183 baseline conditions were significantly associated with subsequent diagnosis of enthesopathy and enthesitis after multiple testing correction (FDR < 0.05) (**Figure 1A**). These associations spanned a wide range of clinical categories, including musculoskeletal, symptom-based, neurological, endocrine/metabolic, circulatory, and respiratory domains. As anticipated, musculoskeletal conditions and other joint-related disorders were prominently associated with future diagnosis of enthesopathy and enthesitis. Osteoarthrosis, other arthropathies, back pain, and intervertebral disc disorders had some of the strongest associations, highlighting shared degenerative and biological mechanisms. These findings are consistent with the localized nature of shoulder pathology and reinforce their clinical relevance. However, beyond these expected associations, the analysis also revealed significant comorbidities across diverse systemic domains. For example, hypertension, asthma, gastritis and duodenitis, and nonspecific chest pain were enriched among incident cases. These findings suggest that enthesopathy and enthesitis may arise within broader physiological contexts, potentially reflecting contributions from vascular, metabolic, inflammatory, or neurogenic pathways. Although a subset of baseline conditions showed statistically significant odds ratios less than 1, indicating a potential inverse association with future disease onset, our analysis primarily focused on comorbidities with odds ratios greater than 1, which reflect conditions enriched in incident cases. This decision aligns with our enrichment-based framework, aimed at uncovering risk-enhancing multimorbidity patterns. This diversity in comorbidity patterns underscores the need for data-driven approaches to identify clinically meaningful endotypes within tendinopathy populations.

**Figure 1.**
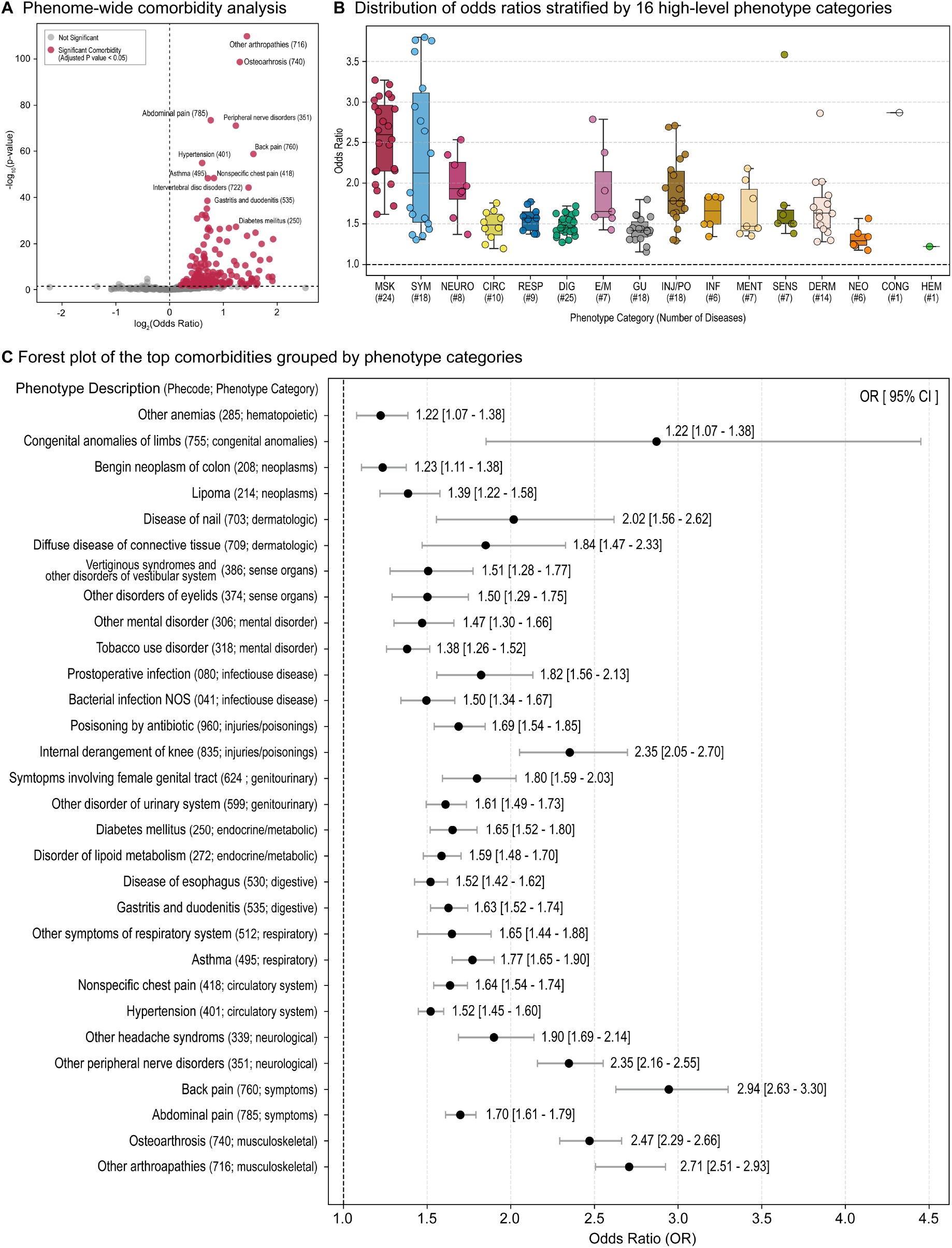
Phenome-wide comorbidity analysis of enthesopathy and enthesitis. (A) Volcano plot of odds ratios and −log_10_(p-values) for 434 baseline conditions; red points indicate FDR-significant associations. (B) Boxplots of odds ratios across 16 phenotype categories highlight strongest enrichment in musculoskeletal, symptom, and neurological domains. (C) Forest plot of top comorbidities per category (OR ≥ 1, FDR < 0.05) with 95% confidence intervals. Results reveal both expected joint-related risks and systemic associations, supporting the need for endotype stratification.

To contextualize these associations across high-level clinical categories, we summarized the distribution of odds ratios for all 16 phenotype domains (**Figure 1B**). Musculoskeletal (MSK), symptom-based (SYM), and neurological (NEURO) conditions exhibited the highest median odds ratios and the widest interquartile ranges, showing both expected localized pathologies and systemic neural involvement. Endocrine/metabolic (E/M), respiratory (RESP), and circulatory (CIRC) categories also showed enrichment, highlighting the multisystem nature of potential risk.

To further highlight representative associations from each clinical category, we present a forest plot of the top comorbidities grouped by domain (**Figure 1C**). For each category, the most significant condition (or top two, if applicable) with an odds ratio ≥ 1 is shown, along with corresponding 95% confidence intervals. As expected, musculoskeletal conditions such as osteoarthrosis and other arthropathies exhibited the highest effect sizes. However, the plot also reveals elevated risk signals from conditions spanning neurological (e.g., headache syndromes, peripheral nerve disorders), respiratory (e.g., asthma), endocrine/metabolic (e.g., diabetes), and gastrointestinal domains (e.g., gastritis and duodenitis). This diversity in comorbidity profiles reinforces the view that tendinopathies and rotator cuff tears may result from multiple, potentially interacting, systemic processes. The cross-domain associations support the need for multidimensional characterization of patient subgroups, motivating downstream network modeling and unsupervised clustering analyses.

### 2.3. Ego Disease Network Analysis Reveals Distinct Endotype Clusters

To better understand the heterogeneity of comorbidities associated with enthesopathy and enthesitis, we constructed a directed disease network centered on the target phenotype using the 183 significant comorbidities identified from the phenome-wide analysis. In this ego-alter structure, the enthesopathy and enthesitis served as the ego node, and each significantly associated condition was represented as an alter node. Directed edges from alter nodes to the ego node indicated significant baseline-to-incident associations.

To capture higher-order structure among comorbidities, we extended the network by estimating pairwise associations between alter nodes using the same odds ratio framework. These alter–alter relationships were modeled as undirected edges, reflecting symmetric comorbidity co-occurrence rather than temporally ordered dependencies. This resulted in a weighted adjacency matrix representing the strength of co-occurrence across all comorbid conditions. We applied Uniform Manifold Approximation and Projection (UMAP) for dimensionality reduction and then performed clustering using the density-based algorithm DBSCAN^17,18^. This unsupervised learning approach revealed eight distinct clusters, each representing a candidate comorbidity-driven subgroup (Figure 2A). Each cluster was annotated based on dominant clinical categories and representative phenotypes (Figure 2B). The following are brief interpretations for each cluster:

**Figure 2.**
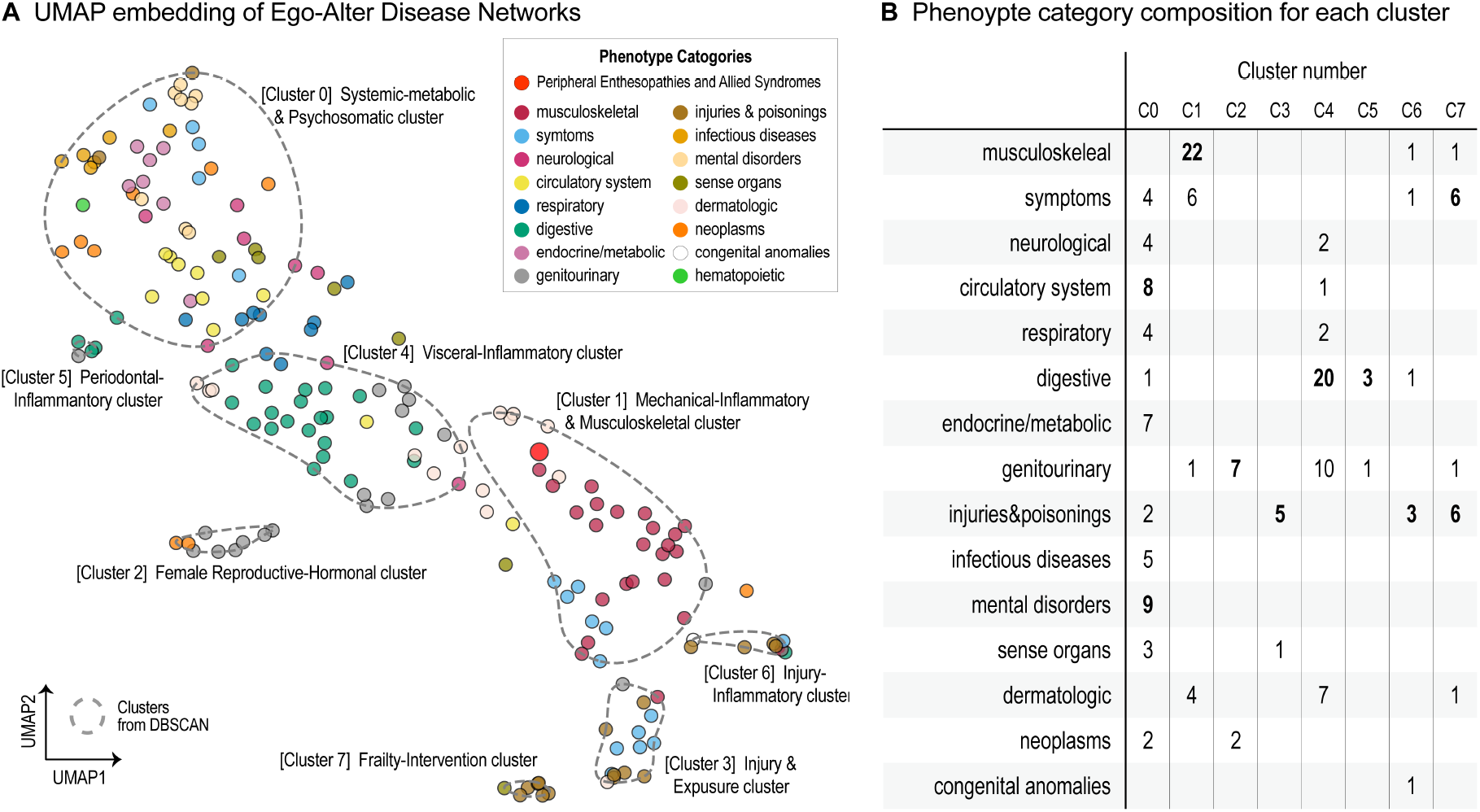
Data-driven comorbidity clusters reveal clinically distinct phenotypic subgroups of enthesopathy and enthesitis. (A) UMAP embedding of ego disease networks constructed from significant baseline comorbidities associated with incident case for target phenotype. Each dot represents a comorbidity, colored by high-level phenotype category. Clustering via DBSCAN identified eight discrete clusters, annotated based on dominant phenotypic domains. (B) Heatmap summarizing the distribution of phenotype categories across clusters. Cluster compositions reveal distinct clinical signatures, including systemic-metabolic (Cluster 0), musculoskeletal-inflammatory (Cluster 1), female reproductive-hormonal (Cluster 2), injury-related (Cluster 3), visceral-inflammatory (Cluster 4), periodontal-inflammatory (Cluster 5), infection-injury (Cluster 6), and frailty-intervention–related (Cluster 7) profiles.

- Cluster 0: characterized by a broad range of metabolic, cardiovascular, and psychiatric comorbidities, including diabetes, hypertension, obesity, hypothyroidism, and depression. This composition suggests a Metabolic– Psychosomatic profile, likely reflecting multisystem dysregulation involving metabolic, vascular, and neuropsychiatric axes ^19^.
- Cluster 1: included the target phenotype of enthesopathy and enthesitis, was enriched for musculoskeletal and autoimmune inflammatory conditions such as osteoarthrosis, spinal disorders, rheumatoid arthritis, and psoriasis. This pattern is consistent with a Mechanical–Inflammatory profile involving degenerative and immune-mediated joint and soft tissue pathologies^3,4,11^.
- Cluster 2: defined by female reproductive and endocrine conditions, including endometriosis, menstrual abnormalities, and benign breast disorders, indicating a Female Reproductive–Hormonal profile. The comorbidity burden suggests hormonal modulation and gynecologic inflammation as potential contributors^20^.
- Cluster 3: injury-related diagnoses such as fractures, contusions, superficial injuries, and poisoning events, indicating an Injury & Exposure profile. This may reflect pathways involving acute physical insult, drug-related toxicity, and downstream tissue damage.
- Cluster 4: a high burden of gastrointestinal, genitourinary, dermatologic, and neuropathic disorders, including irritable bowel syndrome, urinary tract infections, chronic skin ulcers, and peripheral nerve disorders. These features support a Visceral–Inflammatory profile, potentially driven by systemic inflammation, mucosal barrier dysfunction, and autonomic dysregulation.
- Cluster 5: primarily consisted of dental and periodontal diseases, which have been associated with systemic inflammatory responses and musculoskeletal sequelae. This defines a Periodontal–Inflammatory profile, linking chronic oral inflammation to systemic immune activation ^21^.
- Cluster 6: included musculoskeletal infections (e.g., osteomyelitis), congenital limb anomalies, and dislocations, as well as poisoning by psychotropic agents, suggesting an Injury–Inflammation profile characterized by infectious, developmental, and trauma-related factors.
- Cluster 7: dominated by frailty-associated symptoms (e.g., fatigue, syncope), osteoporosis, and complications related to implanted medical devices, consistent with an Aging–Intervention–Related profile involving impaired physiological reserve and heightened vulnerability to iatrogenic events.

This unsupervised learning approach revealed eight distinct clusters, each representing a candidate comorbidity-driven subgroup (Figure 2A). In addition, a small subset of comorbidities did not fall into any cluster based on DBSCAN’s density criteria and were designated as outliers. These outlier traits—positioned between clusters such as Cluster 0 and Cluster 4 or Cluster 4 and Cluster 1—may reflect transitional or mixed phenotypes not strongly affiliated with a single dominant comorbidity group. Each cluster was annotated based on dominant clinical categories and representative phenotypes (Figure 2B).

To enhance clinical interpretability and biological coherence, we consolidated the eight data-driven comorbidity clusters into four unified endotypes based on overlapping pathophysiological mechanisms and phenotypic domains (Table 1). The Metabolic–Psychosomatic Endotype (Cluster 0) encompassed conditions such as obesity, diabetes, hypertension, and psychiatric disorders, reflecting systemic metabolic dysregulation and psychosocial burden. The Inflammatory–Multisystem Endotype (Clusters 1, 2, 4, and 5) integrated a diverse yet immunologically aligned spectrum of conditions, including autoimmune musculoskeletal diseases (Cluster 1), reproductive tract inflammation (Cluster 2), gastrointestinal and dermatologic disorders (Cluster 4), and chronic periodontal disease (Cluster 5). While these clusters represent clinically distinct organ systems, they share common immune-related mechanisms and were located in close proximity within the UMAP embedding space. This spatial contiguity, combined with overlapping inflammatory themes, motivated their grouping into a broader multisystem inflammatory endotype. The Mechanical–Injury–Driven Endotype (Clusters 3 and 6) was defined by trauma-related phenotypes, such as fractures, dislocations, musculoskeletal infections, and tissue damage, consistent with pathways involving biomechanical stress and post-injury inflammation. Finally, the Aging–Intervention–Related Endotype (Cluster 7) comprised of features of physiological frailty, bone fragility, and complications associated with medical devices or procedures, representing a population with reduced resilience and increased susceptibility to treatment-related adverse outcomes. These composite groupings provide a clinically coherent and biologically plausible framework for understanding the diverse systemic profiles associated with rotator cuff pathology and tendon disorders. They offer a foundation for endotype-based stratification and may inform precision risk modeling and intervention strategies in future studies.

**Table 1.**
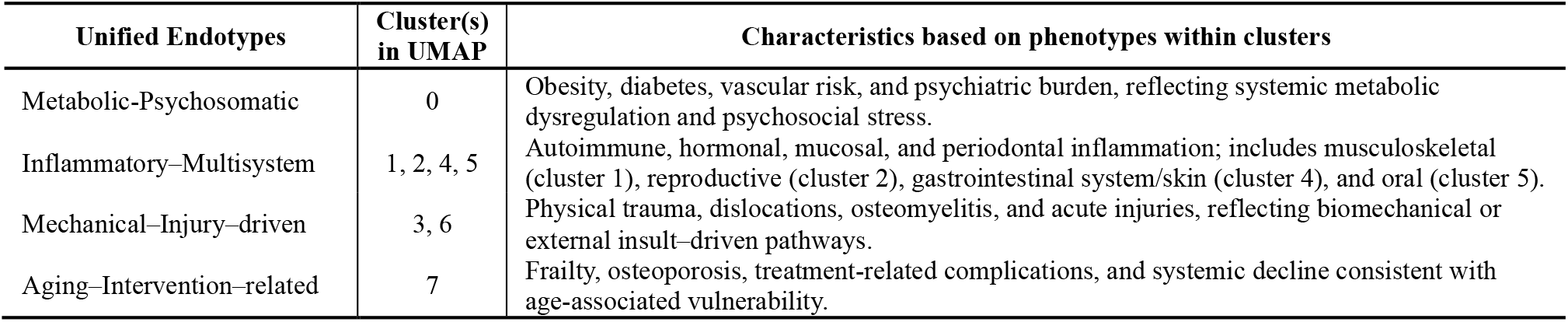
Unified Endotypes dervied from comorbidity cluster groupings.

### 2.4. Metapath-Based Network Analysis Reveals Common Disease Trajectories

To elucidate potential multimorbidity patterns leading to the development of rotator cuff pathology and tendon disorders, we performed a metapath-based network analysis using the constructed ego disease networks^22-24^. Each node in this network represents a disease phenotype, and edges represent statistically significant comorbidity associations derived from baseline-to-incident transitions observed in longitudinal EHR data. A metapath was defined as a directed sequence of one to three disease phenotypes that terminate in the target phenotype (Phecode 726: Peripheral enthesopathies and allied syndromes). Although the source data were not explicitly time-resolved for each pairwise link, the edge directions were based on the temporal ordering of diagnoses relative to baseline, and edge weights reflected odds ratio–based comorbidity strength. For each path, we computed a cumulative metapath score by combining the association strengths along the path, prioritizing trajectories with both statistical support and temporal plausibility. We systematically enumerated and ranked all 1-, 2-, and 3-step metapaths terminating at the target phenotype. The top-ranking 3-step paths were visualized in a layered Sankey-style diagram (**Figure 3)** with nodes color-coded by clinical category and arranged by their sequential position (source → intermediate → target). This visualization integrates both the comorbidity network structure and the cumulative association paths leading to the disease endpoint. We prioritized these trajectories based on their cumulative metapath scores, which aggregated the odds ratio–based association strengths of each sequential link along the path. While individual edges were statistically significant in the initial comorbidity analysis (FDR < 0.05), the full 3-step paths were not evaluated using formal path-level p-values. Instead, the top-ranked metapaths were selected based on their total composite scores and interpreted for biological plausibility and clinical relevance. Several clinically coherent trajectories emerged:

**Figure 3.**
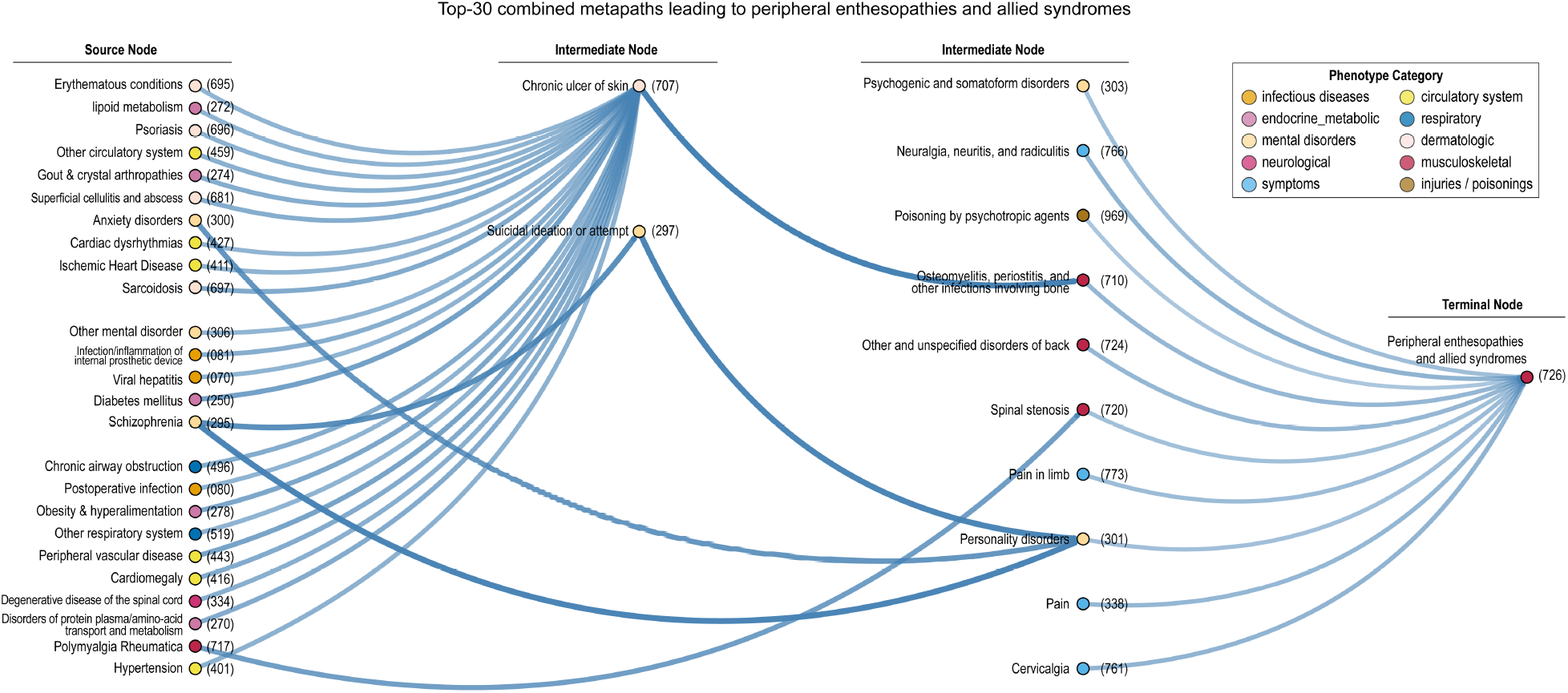
Top-ranked metapaths reveal multimorbidity trajectories culminating in enthesopathy and enthesitis. Directed disease paths of maximum length 3 terminating at Phecode 726 are visualized as a layered network. Source nodes (left), intermediate nodes (middle), and the terminal node (right) are color-coded by clinical category. Edge thickness reflects the strength of pairwise comorbidity associations. Prominent trajectories include metabolic-infectious-musculoskeletal cascades (e.g., diabetes → chronic ulcer of skin → osteomyelitis), inflammatory skin-to-joint sequences, neuropsychiatric-somatic transitions, and vascular-mechanical progressions. This metapath framework highlights plausible clinical chains that may mediate or precede tendon pathology.

- Metabolic-Infectious-Musculoskeletal Cascade: A prominent path began with diabetes mellitus (Phecode 250), progressed through chronic ulcer of skin (Phecode 707) and osteomyelitis (Phecode 710), culminating in enthesopathy and enthesitis. This sequence reflects a well-characterized complication chain involving impaired wound healing and deep tissue infection, often observed in metabolic syndrome.
- Obesity-Vascular-Spinal Progression: Another trajectory traced obesity and hyperalimentation (Phecode 278) to hypertension (Phecode 401), and subsequently to degenerative spinal disease (Phecode 334), indicating a vascular–mechanical route from metabolic dysfunction to musculoskeletal degeneration.
- Inflammatory Skin-to-Joint Sequence: A representative immuno-inflammatory route began with psoriasis (Phecode 696), followed by superficial cellulitis and abscess (Phecode 681), and ended with spinal stenosis (Phecode 720). This pattern underscores systemic inflammatory activation that extends from skin to deeper musculoskeletal structures.
- Neuropsychiatric-Somatic Bridge: One highly interconnected path followed schizophrenia (Phecode 295) → suicidal ideation (Phecode 297) → somatoform disorders (Phecode 303) → pain syndromes, illustrating a psychosomatic progression with downstream somatic complaints.
- Pain-Focused Progression: A frequently observed trajectory began with anxiety disorders (Phecode 300), followed by joint pain (Phecode 773), and culminating in cervicalgia (Phecode 761). This pattern suggests a temporal sequence from mental health burden to progressive regional musculoskeletal pain, consistent with psychosomatic and stress-related pain pathways.

While 1-step metapaths (i.e., direct links from a single baseline comorbidity to enthesopathy and enthesitis) were also included in our analysis, the 3-step sequences provided deeper insight into potential mechanistic chains and disease progression patterns that precede musculoskeletal deterioration. These multi-hop pathways revealed how combinations of metabolic, inflammatory, psychosocial, and mechanical factors may converge over time to increase an individual’s risk for tendon pathology. Altogether, this metapath-based framework supports the observation that rotator cuff pathology and tendon disorders often arise as a downstream manifestation of multi-domain, temporally structured comorbidity patterns. By identifying these intermediate diagnostic routes, this approach enhances interpretability of disease progression and may inform early intervention strategies targeting upstream conditions.

## 3. Conclusion

In this study, we performed a large-scale, phenome-wide comorbidity analysis of enthesopathy and enthesitis using longitudinal EHR data from over 430,000 individuals in the UK Biobank. By identifying 183 baseline conditions significantly associated with the future onset of peripheral enthesopathies and allied syndromes and modeling their relationships through ego–alter disease networks, we uncovered distinct clusters of comorbidities. These clusters were consolidated into four biologically and clinically coherent endotypes—Metabolic–Psychosomatic, Inflammatory– Multisystem, Mechanical–Injury–Driven, and Aging–Intervention–Related—highlighting the multifactorial nature of tendon-adjacent pathology. Furthermore, metapath-based trajectory analysis revealed multistep diagnostic sequences connecting upstream systemic disturbances to the eventual manifestation of enthesopathy, offering a temporal framework for understanding disease progression. This work demonstrates the utility of data-driven, network-based approaches in uncovering latent systemic patterns within musculoskeletal disease populations. The ego–alter network design, combined with unsupervised clustering and trajectory inference, provides a scalable and interpretable framework for identifying comorbidity-driven subtypes that are often obscured in conventional risk modeling. Importantly, the identified endotypes were not limited to musculoskeletal features alone, but captured multisystem domains such as neuropsychiatric disorders, metabolic syndrome, gastrointestinal inflammation, and procedural complications—suggesting that enthesopathy and enthesitis may serve as downstream manifestations of diverse systemic processes.

However, several limitations warrant consideration. First, comorbidity-based endotyping was performed using only diagnostic code data; while comprehensive, this phenotype-centric approach may overlook important biological heterogeneity or subclinical pathophysiology. Second, temporal inferences are constrained by the observational nature of EHRs, which are susceptible to documentation delays, misclassification, and biases in healthcare access or utilization. Third, our focus on incident diagnoses excluded individuals with prevalent or asymptomatic disease, potentially limiting generalizability. Lastly, the endotype assignments were made at the disease level rather than the patient level—future work should incorporate patient-level clustering or risk modeling to enable individualized stratification. To extend these findings, future studies should evaluate the predictive value of endotype membership using prospective or external cohorts, such as All of Us, which includes diverse populations and additional data modalities. The network-derived endotype framework can also be used to inform risk prediction models, patient segmentation, and disease trajectory forecasting. Importantly, incorporating multi-omics data (e.g., genomics, proteomics, metabolomics), lifestyle factors, and environmental exposures may substantially refine endotype definitions and uncover new mechanistic links between systemic physiology and tendon interface disease.

While this framework is primarily designed for discovery, it also lays a foundation for translational application. The identified endotypes may serve as interpretable clinical subgroups that guide future screening, prognostic stratification, or tailoring of therapy. For instance, individuals mapped to a metabolic–psychosomatic endotype might benefit from targeted metabolic risk management or behavioral interventions, whereas those in inflammatory profiles could be candidates for immune-modulating therapies. As electronic health record–linked biobanks become increasingly available, integrating such endotypic classification into risk calculators or clinical decision support tools could enable population-scale risk triage or precision trial enrollment. Further validation in clinical settings and integration with patient-level prediction models will be critical for operationalizing these insights in practice. In conclusion, our study provides a generalizable framework for identifying and interpreting multimorbidity structures in musculoskeletal disease through phenome-scale network modeling. These results underscore the value of integrating comorbidity trajectories and systemic health patterns in understanding enthesopathy and enthesitis, and set the stage for future precision medicine strategies that incorporate biologically informed subtypes into clinical decision-making.

## 4. Methods

### Phenotyping

We utilized hospital inpatient records from the UK Biobank, specifically ICD-10 diagnosis codes extracted from Hospital Episode Statistics (HES), available through Data-Fields 41270 and 41280. For each participant, the baseline was defined using the date of the initial assessment visit (Field 53). By comparing diagnosis dates to this baseline, we categorized each participant as a prevalent case if they had the index disease prior to baseline, and as an incident case if the disease was diagnosed after baseline. ICD-10 codes were mapped to phecodes using the “Phecode Map 1.2 with ICD-9 and ICD-10-CM Codes” available from the PheWAS catalog (https://phewascatalog.org/). To simplify the analysis, we aggregated phecodes to the integer level by truncating the decimal component (e.g., Phecode 250.11 was rolled up to 250). Among the 18,904 unique ICD-10 codes in the HES dataset, we performed the following preprocessing steps: 1) Mapped all ICD-10 codes to phecodes and rolled them up to the top-level integer, 2) Excluded phecodes for which the number of prevalent cases was fewer than 200. After this preprocessing, a total of 453 phecodes were retained for downstream phenome-wide comorbidity analysis.

### Comorbidity Measurement

To identify baseline conditions associated with the future development of peripheral enthesopathies and allied syndromes (Phecode 726), we conducted a phenome-wide comorbidity analysis using hospital diagnosis data from the UK Biobank. For each candidate condition, we tested whether its presence at or before baseline was associated with a higher incidence of phecode 726 during follow-up. We restricted our analysis to individuals without prevalent enthesopathy and enthesitis at baseline (i.e., those whose first diagnosis occurred after the date of initial assessment). For each of the 434 candidate phecodes, we determined whether a participant had a diagnosis prior to baseline and constructed a 2×2 contingency table comparing the presence of the underlying condition against the incidence of enthesopathy and enthesitis^15,25^. We computed odds ratios using Fisher’s exact test, and applied false discovery rate (FDR) correction to control for multiple testing^26^. Phecodes with FDR-adjusted p-values less than 0.05 were considered significantly associated with the future onset of peripheral enthesopathies. This phenome-wide approach identified 183 baseline conditions significantly enriched among incident cases.

### Ego Disease Network Construction

We constructed a directed and weighted disease network to represent the phenome-wide comorbidity structure centered on the index phenotype: peripheral enthesopathies and allied syndromes. This ego–alter network designates the target phenotype as the ego node, and the 183 comorbid conditions significantly associated with its future incidence (FDR < 0.05) as alter nodes ^27,28^. Edges in the network were directed from each alter node to the ego node, reflecting the temporal and statistical structure of the comorbidity analysis—specifically, the predictive association between baseline diagnoses of alter conditions and incident enthesopathy and enthesitis. Edge weights were defined using odds ratios derived from Fisher’s exact tests conducted in the comorbidity analysis, providing a quantitative measure of association strength.

### Comorbidity Clustering with embedded Ego Network

To identify latent structure among the significant comorbidities associated with the target disease, we applied unsupervised clustering to a comorbidity network derived from the ego–network structure. The ego network consisted of one central node representing the target condition (Phecode 726: peripheral enthesopathies and allied syndromes) and 183 significantly associated comorbidities as alter nodes. Directed edges from each alter to the ego node were weighted by odds ratios derived from the phenome-wide comorbidity analysis, reflecting the statistical strength of association between each baseline diagnosis and future development of the target condition. To investigate potential higher-order structure among comorbidities themselves, we constructed a separate undirected alter–alter network, in which each pair of comorbid conditions was connected by an edge weighted by their co-occurrence strength across individuals. These pairwise co-occurrence scores were estimated using the same odds ratio framework as in the ego network, but the ego node (Phecode 726) was excluded from this analysis. The resulting 183 × 183 adjacency matrix captured comorbidity co-occurrence patterns across all alter nodes. We then applied Uniform Manifold Approximation and Projection (UMAP) to embed this matrix into a low-dimensional space while preserving its global and local structure. On the resulting 2D embedding, we applied Density-Based Spatial Clustering of Applications with Noise (DBSCAN) to detect distinct comorbidity clusters. Each cluster was interpreted as a candidate endotype based on shared clinical themes, and annotated using dominant phenotype categories and representative diagnoses.

### Metapath Analysis

To investigate potential multimorbidity trajectories culminating in the target disease, we performed a metapath analysis on the directed ego network. A metapath was defined as a directed sequence of one to three diagnoses, where each edge in the sequence represented a statistically significant comorbidity association leading to the terminal node: Phecode 726 (enthesopathy and enthesitis). This design allowed us to identify both direct (1-step) and indirect (2- or 3-step) diagnostic routes potentially reflective of progressive systemic deterioration. We first enumerated all acyclic directed paths of length 1 to 3 that terminated in the ego node. Each edge in the path corresponded to a significant association from the original comorbidity network, with edge weights defined by odds ratios (ORs) from Fisher’s exact tests. To compute the cumulative score of a given metapath, we aggregated the base-10 logarithm of the odds ratios across each step:

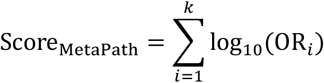

where *k* is the number of steps (1 to 3) in the path. This additive log-scale scoring emphasizes sequences with consistently strong associations while maintaining interpretability across paths of different lengths. Only paths with positive cumulative scores (i.e., all ORs > 1) were retained for downstream ranking and visualization. Top-ranked 3-step metapaths were visualized using a layered Sankey-style flow diagram. Each node was color-coded by its clinical category (e.g., metabolic, neurological), and positioned according to its path role (source, intermediate, or terminal node). This framework highlights clinically coherent multimorbidity trajectories and suggests interpretable diagnostic progressions that may precede enthesopathy and enthesitis.

## Data Availability

We acknowledge all the participants of the UK Biobank. The use of the UK Biobank resource was approved under Application Number 32133.

